# DiNetxify: a Python package for three-dimensional disease network analysis based on electronic health record data

**DOI:** 10.1101/2025.08.19.25333629

**Authors:** Can Hou, Haowen Liu, Viktor H. Ahlqvist, Elisabet Unnur Gisladottir, Yao Yang, Huazhen Yang, Fang Fang, Unnur A. Valdimarsdóttir, Huan Song

## Abstract

The rapid expansion of large-scale electronic health record (EHR) data has underscored the necessity for advanced analytical methods, such as disease network analyses, to comprehensively identify and interpret multimorbidity patterns and disease progression pathways. To overcome existing obstacles associated with performing sophisticated disease network analyses on EHR data, we developed ***DiNetxify***, an open-source Python package implementing our recently introduced three-dimensional (3D) disease network analysis method (https://hzcohort.github.io/DiNetxify/). ***DiNetxify*** provides a dedicated data class for handling various EHR data, comprehensive modular functions for executing complete 3D disease network analyses, and visualization functions for interactive exploration of results. The package is efficient, user-friendly, and optimized for large-scale EHR datasets. It supports diverse study designs, customizable analysis parameters, and parallel computing for enhanced performance. Through two case studies utilizing the UK Biobank data, one investigating disease networks associated with short leukocyte telomere length and the other exploring disease networks in the middle-aged general population, we demonstrated the capability of ***DiNetxify*** to identify meaningful disease clusters and progression patterns consistent with established knowledge while uncovering novel insights. Computationally, the software successfully completed analyses involving cohorts exceeding half a million exposed individuals within 17 hours, using moderate computational resources. We thus anticipate that ***DiNetxify*** can significantly reduce technical barriers to facilitate broader adoption of advanced disease network analysis techniques by different researchers, thereby enhancing the exploration of EHR data to improve the understanding of holistic health dynamics.

## Background

The rapid expansion of population-scale electronic health record (EHR) has created an urgent need for innovative analytical approaches capable of identifying and interpreting the complex correlations across hundreds of diseases documented in millions or even billions of EHR entries. Disease network analyses, encompassing both disease trajectory and comorbidity network analyses, have emerged as powerful computational tools that extend beyond traditional hypothesis-driven studies of single diseases by exploring intricate relationships among multiple disorders^1^. For instance, population-wide disease trajectory analyses have identified numerous disease progression sequences (i.e., disease trajectories) from datasets containing millions of patients, thereby facilitating the discovery of novel disease associations, potential pathways for intervention, and planning of healthcare systems^2,3^. Similarly, comorbidity network analyses, which examine disease co-occurrence irrespective of temporal sequence, have successfully identified meaningful multimorbidity clusters that may reflect shared etiology^4–6^.

Despite these advancements, current disease network methodologies exhibit notable limitations. They often analyze temporal and non-temporal associations separately, possess limited statistical power, and are susceptible to spurious associations due to confounding by concurrent comorbidities. To mitigate these issues, we recently developed a three-dimensional (3D) disease network analysis approach that integrates disease trajectory and comorbidity network analyses^7^. This method incorporates regularized partial correlation networks and advanced 3D visualization techniques, enabling robust identification of disease clusters across diverse EHR datasets collected from multiple countries. Nevertheless, a broader application of disease network analysis remains challenging, as most existing tools and scripts are customized for specific datasets or address only part of the often complex disease network analytical workflow^8–10^. The limited generalizability and lack of standardized, user-friendly software further discourage adoption among researchers without extensive programming skills.

To address these barriers, we developed ***DiNetxify***, an open-source Python package designed to implement the comprehensive 3D disease network analysis approach. ***DiNetxify*** provides an end-to-end analytical solution encompassing EHR data preprocessing, disease network analysis, and interactive visualization. Optimized for large-scale datasets, it features a user-friendly interface that significantly lowers technical barriers, enabling researchers to readily apply sophisticated disease network analysis methods to extensive EHR datasets. We illustrate the capabilities and evaluate the performance of ***DiNetxify*** through two distinct case studies employing different study designs.

## Methods

### Framework of 3D disease network analysis

The 3D disease network analysis implemented in ***DiNetxify*** builds upon our previously published methodology after incorporating enhancements optimized for software implementation^7^. By integrating traditional disease trajectory and comorbidity network analyses using regularized partial correlations, the framework simultaneously captures temporal progression and non-temporal associations within a unified three-dimensional space. The analytical workflow consists of five key steps:

**(1) Phenome-wide association study (PheWAS)**: Initially, diseases significantly associated with an index exposure are identified. Depending on the study design, either Cox regression (for cohort studies) or stratified Cox regression (for matched cohort studies) is applied. Hazard ratios (HRs) are calculated after adjustment for covariates, and by default, diseases exhibiting statistically significant positive associations (q-value<0.05 and HR>1) are retained. In cohort studies with exposed-only design, this step is simplified by filtering diseases based on a minimum number of incident cases, due to the absence of a comparison group.
**(2) Disease pair construction and filtering:** From the retained diseases, we construct all possible disease pairs, then evaluate their comorbidity strength using relative risk (RR, i.e., the probability of observing both conditions in the same individual relative to expectation) and Φ-correlation (i.e., Pearson’s correlations for two binary variables) metrics. By default, we keep only disease pairs with RR>1, Φ-correlation>0, and both q-values<0.05. This screening step reduces the search space for disease trajectory and comorbidity network analyses, since only disease pairs with at least some extend of comorbidity strength are likely to contribute meaningfully to networks or trajectories.
**(3) Disease trajectory analysis:** For each retained disease pair, we test temporal directionality. A binomial test assesses whether disease D1 more often precedes disease D2 than the reverse order. Candidate disease pairs undergo further analysis using a nested case-control design where the controls are selected using the method of incidence density sampling. The earlier disease (D1) is modeled as the exposure, and the later disease (D2) as the outcome in conditional logistic regression analyses. Regularized partial correlation is applied by incorporating all other diseases (excluding D1 & D2) as covariates and using an L1 penalty to shrink irrelevant coefficients toward zero^11^. Optimal penalty weights are selected based on the minimum Akaike Information Criterion (AIC). For computational efficiency in large datasets, we introduced a principal component analysis (PCA)-based approximation, substituting the full regularized partial correlation with principal components derived from other diseases, thereby maintaining analytical accuracy while reducing computational complexity.
**(4) Comorbidity network analysis:** Non-temporal associations are estimated using unconditional logistic regression with the same regularized partial correlation framework, or the PCA based approximation. Following that, the Louvain algorithm partitions the comorbidity network into distinct modules.
**(5) Spatial embedding in 3D space:** Finally, each disease is assigned coordinates in a three-dimensional space. The x-y plane represents module membership derived from the comorbidity network, and the z-axis indicates temporal order derived from the disease trajectory analysis. Reliable disease clusters are thus defined as groups of diseases sharing comorbidity module membership and linked through temporal relationships.

### Implementation of 3D disease network analysis in *DiNetxify*

We implemented the above analytical framework in ***DiNetxify***, an open-source Python package designed for the end-to-end, comprehensive disease network analysis of large-scale EHR data (**Figure 1**). The package is available on GitHub https://github.com/HZcohort/DiNetxify and can be installed via PyPI for use on Linux or Windows computers. Extensive documentation, including detailed API references and step-by-step tutorials using example datasets, is provided on the package website (https://hzcohort.github.io/DiNetxify/), facilitating rapid adoption by researchers with limited Python skills.

**Figure 1.**
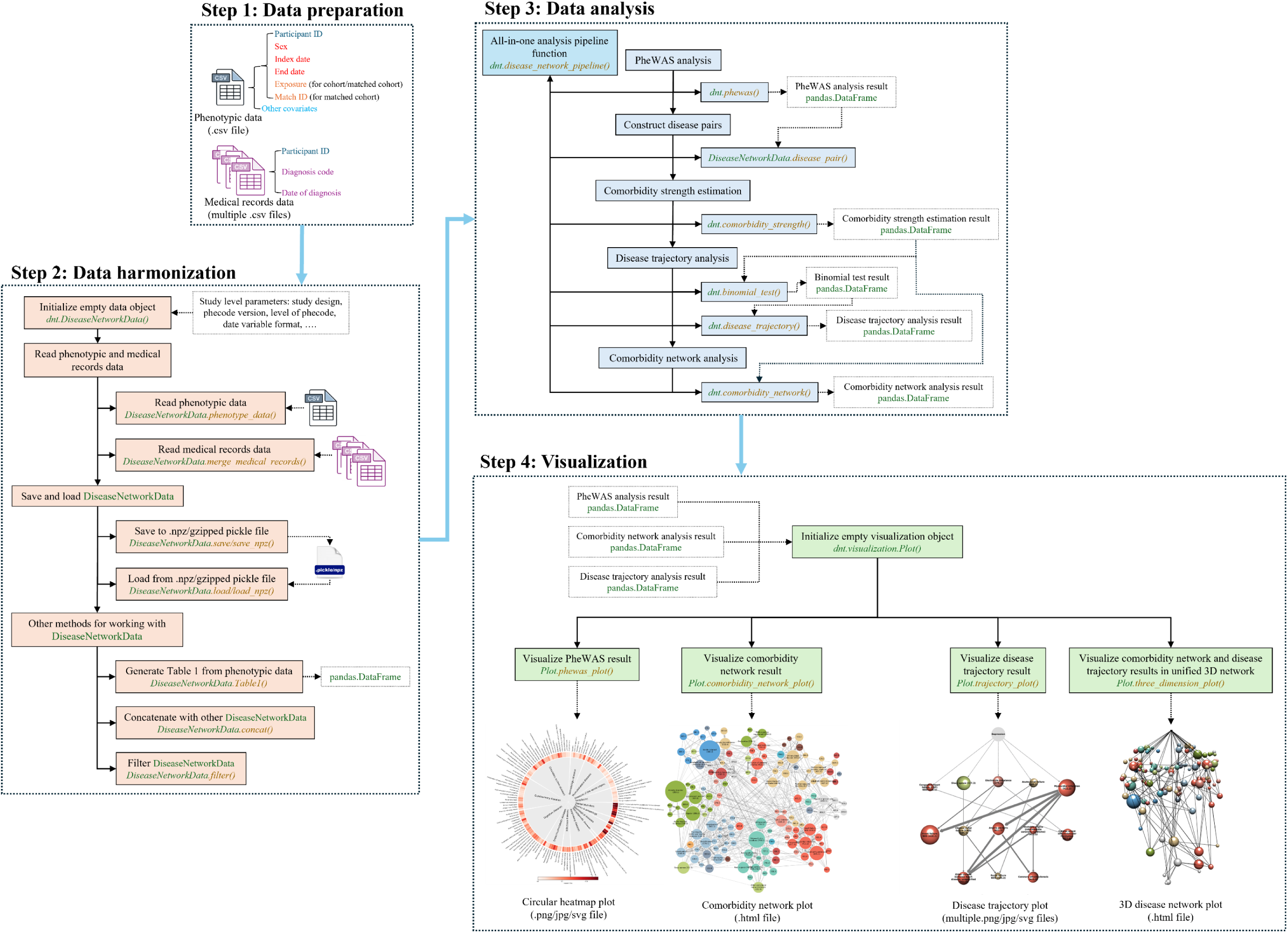
Modular architecture of the DiNetxify package.

#### Dedicated data class

The core of the package architecture is the *DiseaseNetworkData* class, which standardizes and integrates participant-level phenotypic data with record-level EHR data into a unified data structure. This design streamlines data management, optimizes memory use, and enhances data retrieval performance for downstream analyses. Currently, the class supports loading ICD-9 and ICD-10 codes (both WHO and clinical modification versions) from EHR datasets, automatically mapping them to standardized ‘phecodes’^12^, thus eliminating the need for manual diagnosis-code preprocessing. Utilizing ‘phecodes’ also standardizes disease definitions and make full use of their hierarchical structure and associated metadata (such as exclusion criteria and sex-specific codes), enabling accurate and consistent sub-cohort construction. The class supports two ‘phecodes’ levels: level 1 corresponds to 3-digit ICD-10 codes, covering 585 medical conditions, while level 2 corresponds to 4-digit ICD-10 codes, covering 1,257 medical conditions. Level 2 provides greater granularity, suitable for large-scale studies, whereas level 1 is recommended for smaller studies to ensure sufficient statistical power.

#### Analysis module

***DiNetxify*** implements a comprehensive analytical pipeline through modular functions that can be executed either sequentially or automatically via an integrated pipeline function operating on the *DiseaseNetworkData* class. Core analytical functions include *phewas* for phenome-wide association analysis, *comorbidity_strength* for evaluating disease pair associations, *binomial_test* for temporal ordering assessment, *comorbidity_network* for analyzing non-temporal disease networks, and *disease_trajectory* for temporal disease trajectory analysis. Each function allows extensive parameter customization, supports multiple statistical correction methods, and includes parallel processing capabilities. This modular design provides flexibility for diverse research applications and facilitates integration with other analytical toolkits.

#### Visualization module

Visualization capabilities are implemented through the *Plot* class, which receives outputs from analysis functions and offers comprehensive methods for exploring and presenting disease network results. The *Plot* class enables visualization of PheWAS outcomes via circular heatmap plots, comorbidity and disease trajectory networks via network graphs, and the final 3D disease network using interactive 3D network plots. Visualization outputs can be exported in multiple formats suitable for academic publication and presentation purposes.

#### Optimization for large-scale EHR analysis

To enhance the performance of large-scale EHR analyses, the package integrates optimization strategies. Multiprocessing support enables parallel computation on systems with multiple CPU cores, substantially reducing execution times for computationally intensive tasks. Additionally, chunk-wise data processing handles memory-intensive operations by dividing datasets into manageable segments, thus preventing memory overflow issues in large-scale datasets. Based on our experience, medium-sized studies, with up to around 50,000 exposed participants, can be run on standard office laptops, such as those with an Intel Core i5 processor (4 cores) and 16 GB RAM. However, for larger or more complex datasets, high-performance computing resources are recommended to ensure efficient processing.

### Case studies using UK Biobank data

To showcase the utility and performance of ***DiNetxify***, we conducted two case studies using data from the UK Biobank, a community-based prospective cohort of approximately 500,000 middle-aged adults recruited across the UK^13^. Baseline demographics, lifestyle information, and blood biomarkers were collected at enrollment through comprehensive questionnaires and biological sampling. Additionally, EHR data for all participants were regularly linked to multiple national health registries across England, Wales, and Scotland from 1997 to 2022, including mortality data and inpatient hospital diagnoses, all recorded using ICD-9 or ICD-10 codes.

We constructed two separate cohorts from the UK Biobank to represent distinct study designs: a cohort study comparing individuals with short leukocyte telomere length (LTL) (as the exposed group) against all others (as unexposed), and an exposed-only cohort study that included all UK Biobank participants as ‘exposed’ (**Supplementary Figure 1**). Baseline demographic and lifestyle data collected for each participant included age, sex, socioeconomic indicators (Townsend deprivation index and household income), body mass index (BMI), smoking status, alcohol consumption, physical activity, and dietary patterns. Participants were prospectively followed from recruit until 2022, with disease and death outcomes ascertained from linked EHR data.

To assess the performance of ***DiNetxify*** in these cohorts, we executed the complete analytical pipeline using 10, 20, and 40 CPU cores (Intel Xeon Gold 6248 CPU, 2.50 GHz), and compared the total computational time required. Additionally, we performed comorbidity network and disease trajectory analyses using both the PCA-based partial correlation approximation and the original regularized partial correlation approach, comparing computational efficiency and analytical consistency between the two methods.

## Results

### Case study 1: disease network associated with short leukocyte telomere length

We first applied ***DiNetxify*** to analyze the disease network associated with short LTL using data from the UK Biobank. The study cohort included 472,277 individuals with available LTL measurements at recruitment (baseline). Among them, 118,070 individuals were classified as having short LTL (exposed group, defined as individuals within the lowest quartile), while the remaining 354,207 individuals comprised the unexposed group. The median age at baseline was 60.9 years (IQR: 53.9-65.3) in the exposed group and 57.3 years (IQR: 49.7-63.0) in the unexposed group, including 48.25% and 56.18% female participants, respectively.

In the PheWAS analysis, we investigated 1,019 level-2 medical conditions after excluding pregnancy complications, congenital anomalies, injuries and poisonings, and symptom-based conditions. We identified 134 medical conditions with elevated risk among individuals with short LTL (HR>1, q-value<0.05, **Supplementary Figure 2**), after requiring at least 500 incident cases within the exposed group. The strongest associations were observed for post-inflammatory pulmonary fibrosis (HR: 1.92; 95% CI: 1.79-2.06), thrombocytopenia (HR: 1.35; 95% CI: 1.27-1.44), liver abscess and sequelae of chronic liver disease (HR: 1.35; 95% CI: 1.24-1.47), and bronchopneumonia and lung abscess (HR: 1.33; 95% CI: 1.21-1.45). The subsequent comorbidity network analysis identified 1,114 statistically significant non-temporal disease associations among these conditions, partitioning the network into eight distinct modules (**Supplementary Figure 3**). At the same time, the disease trajectory analysis, requiring temporal relationships within a window of one month to five years, yielded 330 statistically significant temporal disease pairs. Integrating these temporal and non-temporal associations resulted in a comprehensive 3D disease network for short LTL (**Figure 2A**, interactive 3D plot can be found at https://hzcohort.github.io/DiNetxify/LTL_RPCN.html). The combined 3D network highlighted eight robust disease clusters (**Figure 2B**), showing major pathways by which short telomere length contributes to an increased disease burden across multiple physiological systems. These clusters include cerebrovascular and neuropsychiatric diseases cluster (cluster 1), hepatobiliary diseases cluster (cluster 2), musculoskeletal diseases cluster (cluster 3), vascular and ocular diseases cluster (cluster 4), gastrointestinal and nutritional diseases cluster (cluster 5), genitourinary and infectious diseases cluster (cluster 6), cardiovascular diseases cluster (cluster 7) and respiratory diseases cluster (cluster 8).

**Figure 2.**
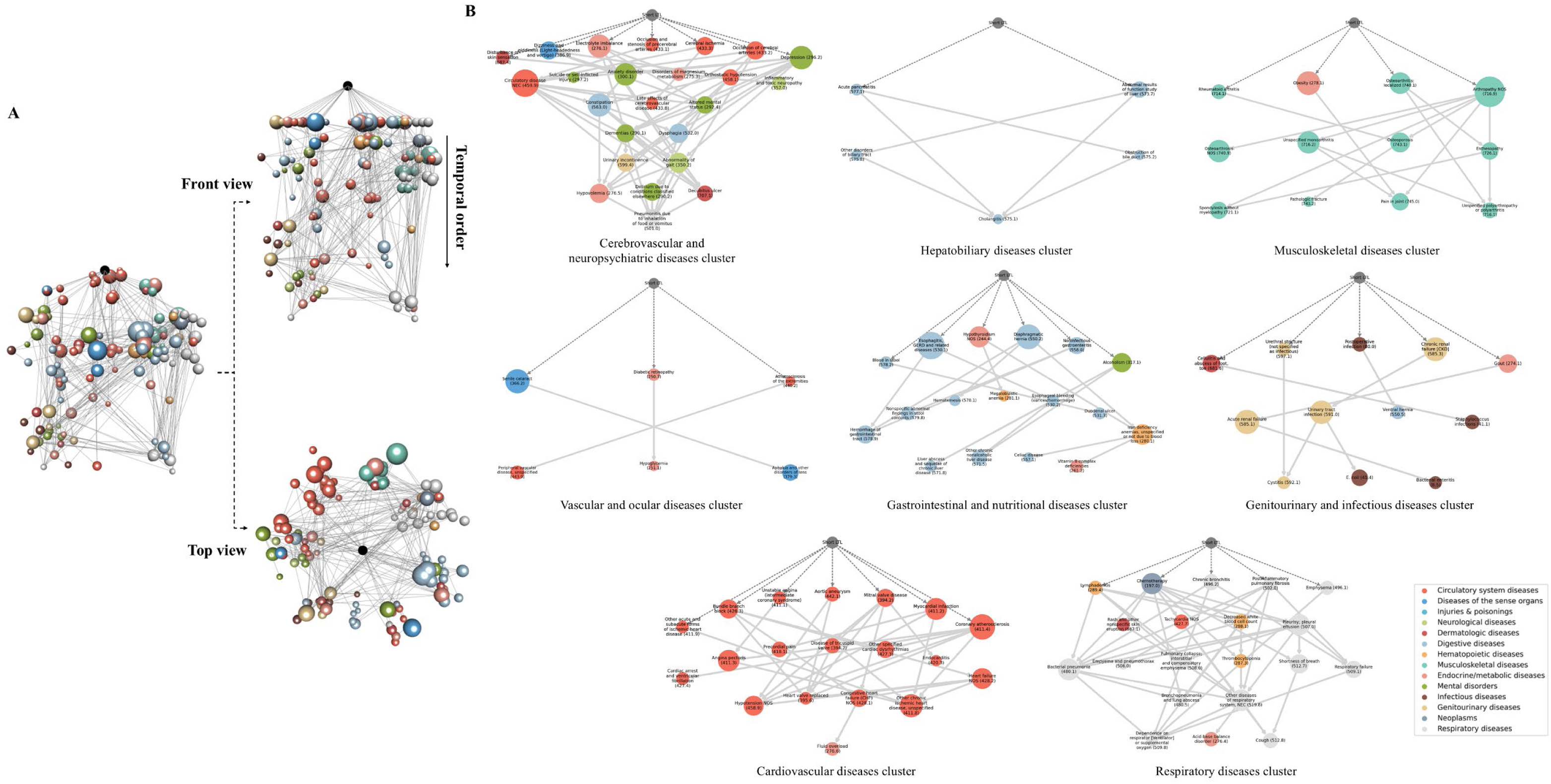
Three-dimensional disease network and the reliable disease clusters associated with short leukocyte telomere length. Visualization of the disease network associated with short leukocyte telomere length. Nodes represent medical conditions defined by level 2 ‘phecodes’, sized by incidence and colored by disease category. **(A)** Integrated three-dimensional network plot combining the comorbidity network (viewed from above) and temporal disease trajectory network (viewed from the side), interactive version at https://hzcohort.github.io/DiNetxify/LTL_RPCN.html. **(B)** Eight robust disease clusters revealed through both comorbidity network and disease trajectory analyses.

We also evaluated computational performance and scalability of ***DiNetxify*** using this cohort. The complete analysis pipeline, including data harmonization and disease network analyses using the regularized partial correlation approach, required approximately 13 hours on a high-performance server equipped with 40 CPU cores and 100 GB of memory (**Figure 3**). On a smaller server with 10 CPU cores, the analysis was completed within approximately 35 hours, demonstrating the feasibility of ***DiNetxify*** for processing large-scale cohorts (approximately 16 million diagnosis records from over 470,000 individuals). Furthermore, we repeated the disease network analysis using the PCA-based partial correlation approximation approach, which significantly reduced computation time to under 1 hour on the 40-core server, indicating approximately a 13-fold improvement in speed (**Figure 3**). When comparing results from the approximation and the regularized partial correlation methods, we observed very high consistency in the estimated beta coefficients for both temporal and non-temporal disease associations (Pearson correlation of 0.985 and 0.860, respectively; **Supplementary Figure 4**). The robust disease clusters identified by the PCA-based approach closely matched those generated by the original method (**Supplementary Figure 5**). These findings indicate that the PCA-based partial correlation approximation is a reliable alternative that provides considerable computational savings with minimal loss in analytical precision, particularly suitable for extremely large datasets or settings with limited computational resources.

**Figure 3.**
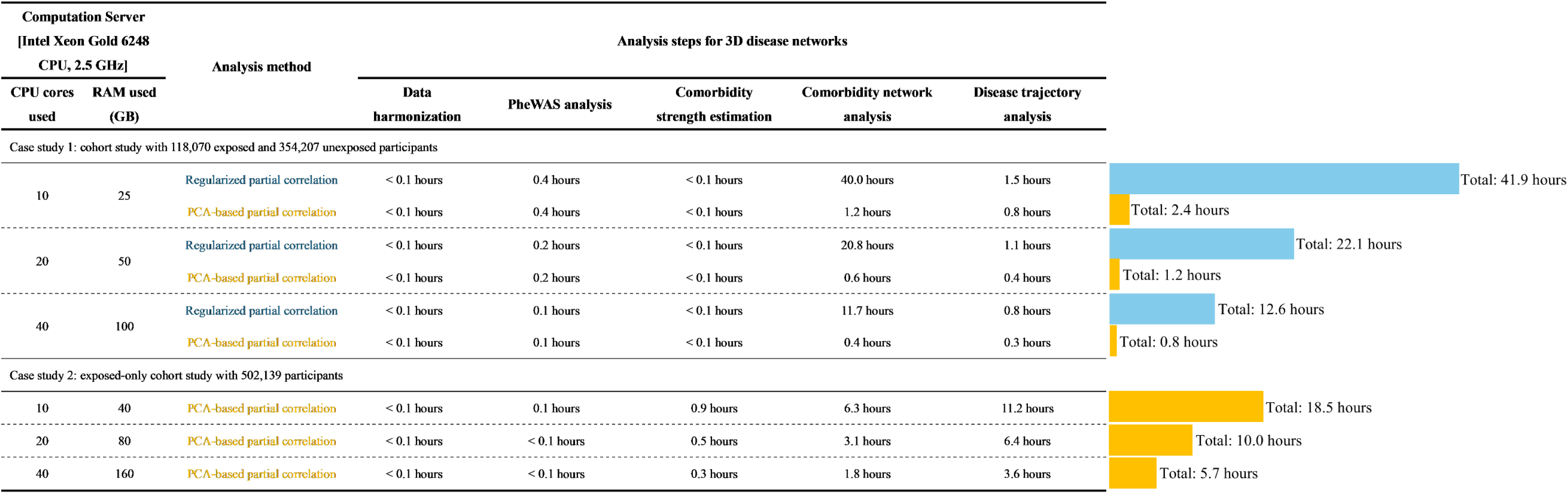
Computational performance of DiNetxify in two UK Biobank case studies. Wall clock runtimes for the 3D disease network workflow across two UK Biobank based case studies. Analyses ran on an Intel Xeon Gold 6248 server, 2.5 GHz, with 10, 20, or 40 CPU cores and 25 to 160 GB RAM allocated. Columns report step specific runtimes, including data harmonization, PheWAS, comorbidity strength estimation, comorbidity network analysis, disease trajectory analysis, and the bars at right show total runtime per configuration. Blue indicates regularized partial correlation approach, yellow indicates the principal component analysis (PCA) based approximation approach.

### Case study 2: disease network in a middle-aged general population

For the second case study, we applied ***DiNetxify*** to an exposed-only cohort from the UK Biobank to investigate typical multimorbidity structures within the middle-aged general population. We included all participants who were aged approximately 40 to 69 years at baseline, resulting in a cohort of 502,139 individuals after excluding those withdrawn from the study. The median age at baseline was 58.3 years (IQR: 50.6–63.7), including 54.40% females. We focused on the same set of 1,019 level-2 medical conditions as in the first case study.

Due to the exposed-only cohort design, the PheWAS analysis filtered for medical conditions with at least 2,500 incident cases instead of based on the difference between exposed and un-exposed as under the first case study, yielding 233 conditions for subsequent network analysis (**Supplementary Figure 6**). Given the substantial cohort size (approximately 0.5 million individuals) and number of medical conditions included, we opted to use the PCA-based partial correlation network approach considering computational efficiency. In the comorbidity network analysis, we identified 4,149 significant non-temporal disease pairs among these 233 conditions. Community detection partitioned this network into nine modules characterized by frequent co-occurrence (**Supplementary Figure 7**). The disease trajectory analysis identified 1,923 significant temporal associations. Combining the non-temporal and temporal associations allowed us to construct a comprehensive 3D disease network (**Figure 4A**, interactive 3D plot can be found at https://hzcohort.github.io/DiNetxify/general_PCA.html), from which we defined nine major disease clusters reflecting common multimorbidity patterns within this population (**Figure 4B**). These clusters included cardiovascular diseases cluster (cluster 1), ocular diseases cluster (cluster 2), oncology and metabolic diseases cluster (cluster 3), gynecologic diseases cluster (cluster 4), gastrointestinal and hepatobiliary diseases cluster (cluster 5), neurological and cerebrovascular diseases cluster (cluster 6), genitourinary and infectious diseases cluster (cluster 7), respiratory diseases cluster (cluster 8), and musculoskeletal and connective tissue diseases cluster (cluster 9).

**Figure 4:**
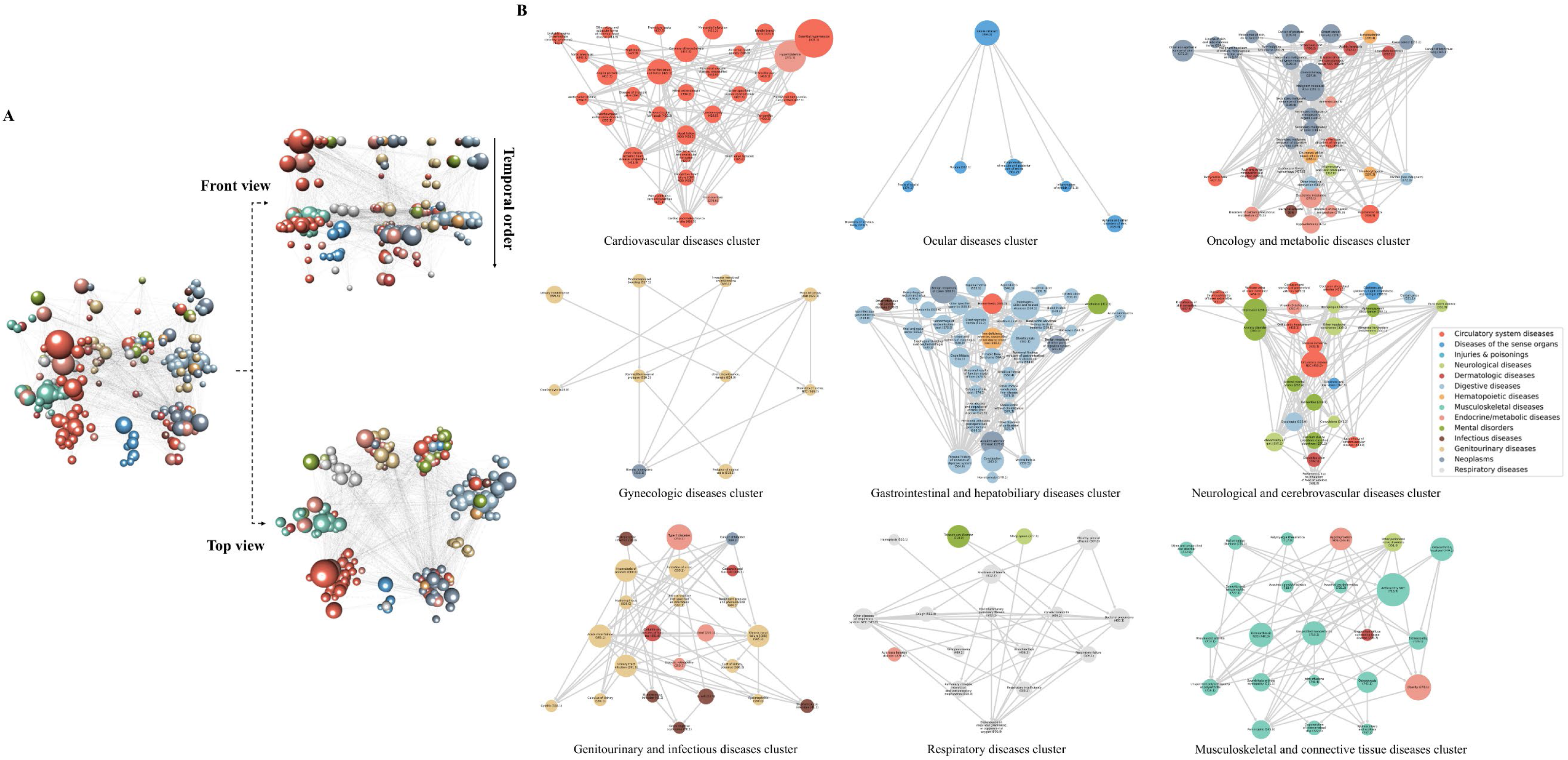
Three-dimensional disease network and the reliable disease clusters in a middle-aged general population. Visualization of the disease network associated in a middle-aged general population. Nodes represent medical conditions defined by level 2 ‘phecodes’, sized by incidence and colored by disease category. **(A)** Integrated three-dimensional network plot combining the comorbidity network (viewed from above) and temporal disease trajectory network (viewed from the side), interactive version at https://hzcohort.github.io/DiNetxify/general_PCA.html**. (B)** Nine robust disease clusters revealed through both comorbidity network and disease trajectory analyses.

The full analytical pipeline was completed in approximately six hours using a server with 40 CPU cores and 160 GB of memory, and roughly 17 hours on a server with 10 CPU cores and 40 GB of memory (**Figure 3**). Although the runtime was longer compared to that of the first case study as the latter had a smaller number of exposed individuals, it runtime remains feasible for a comprehensive, one-time analysis on datasets comprising over half a million individuals.

## Discussion

In this study, we introduced ***DiNetxify***, an open-source Python package implementing our recently proposed 3D disease network analysis approach. ***DiNetxify*** provides comprehensive insights into the multimorbidity patterns and disease progression pathways using EHR data. ***DiNetxify*** emphasizes user-friendliness by integrating the entire analytical pipeline, from data harmonization and analysis to visualization, into a single streamlined Python package. The package is flexible, supporting various study designs and EHR data formats, and is computationally efficient through parallel processing and algorithmic optimizations. Through two case studies with distinct study designs, we demonstrated the capability of ***DiNetxify*** to elucidate complex disease networks following specific exposures or within the general population. These examples also illustrated that large-scale EHR datasets, encompassing cohorts of up to half a million individuals and hundreds of diseases, can be efficiently analyzed using modest computational resources. By releasing ***DiNetxify***, we aim to facilitate a broader adoption of advanced disease network analysis methods among researchers across diverse medical disciplines, thereby promoting high-quality, replicable, hypothesis-free studies using EHR datasets.

Existing software tools for analyzing disease trajectories or comorbidities have notable limitations, such as reliance on uncommon programming languages, incomplete analytical capabilities, or design constraints (i.e., application for specific EHR data), restricting therefore their wider usage. For instance, PTRA, a software tool implemented in the Go language, supports only conventional disease trajectory analysis tailored specifically to TriNetX data^8^. Similarly, LUKB DT, an R Shiny application, facilitates standard disease trajectory analyses but is exclusively designed for the UK Biobank dataset^10^. In contrast, ***DiNetxify*** distinguishes itself by fully supporting advanced 3D disease network analysis, which provides more reliable and interpretable insights. The package encompasses the entire workflow, from raw EHR data preprocessing to sophisticated analyses and interactive visualizations, in a single integrated toolkit. Its flexible architecture supports analysis of diverse EHR datasets with minimal preprocessing requirements and accommodates both conventional and advanced analytical methods through modular design and customizable parameters. Researchers can selectively run specific parts of the analytical pipeline (e.g., PheWAS using Cox regression) or opt for analyses without partial correlation adjustments, according to specific research needs. Additionally, its open-source, Python-based structure facilitates broad usage and user contributed improvements. Despite these strengths, several future enhancements are planned for ***DiNetxify***. including disease network-based risk prediction, patient stratification, the identification of disease networks preceding specific exposures (i.e., the inverse of the current approach), causally informed trajectories and networks, and expansions beyond ‘phecodes’. Such future development may position ***DiNetxify*** as an even more comprehensive analytical toolkit for exploring EHR data.

We showcased the utility of ***DiNetxify*** through two real-world case studies representing common scenarios for disease network analyses. In both cases, ***DiNetxify*** demonstrated strong computational performance, largely attributable to optimizations such as the dedicated data class structure, multiprocessing capabilities, and our PCA-based approximation method. Notably, using the PCA-based method allowed completion of the entire analytical pipeline in approximately 5.5 hours for a cohort with over half a million exposed individuals. The comparison of our results with prior findings provided additional validation. For instance, in the short LTL cohort (case study 1), the identified clusters included cardiovascular, respiratory, cerebrovascular and neuropsychiatric, and musculoskeletal diseases, further supporting established associations between short LTL and increased risk for age-related diseases^14–18^ as well as key pathologies related to telomere dysfunction^19^. Similarly, the identified disease clusters within the middle-aged general population cohort (case study 2) are consistent with known multimorbidity patterns characteristic of this critical life stage^20–22^, highlighting prominent drivers of morbidity and mortality among middle-aged adults. The results also provide novel insights. For example, within the gastrointestinal and nutritional disease cluster associated with short LTL, we identified a temporal progression from hypothyroidism to nutritional-deficiency disorders among individuals with short LTL, a connection not previously recognized. Furthermore, comparing the disease networks of individuals with short LTL to those of the general aging population revealed increased cross-system connectivity in short LTL-associated disease networks, illustrating how accelerated biological aging can fundamentally reshape disease clustering.

While ***DiNetxify*** represents a powerful new tool for disease network analysis using EHR data, several limitations should be acknowledged. First, as with all observational analyses relying on EHR data, results are associative and cannot directly establish causality. Although we have attempted to mitigate this limitation by adjustment for known confounders (e.g., age, sex, socioeconomic status) and other diseases within the network, residual confounding due to unmeasured or unknown factors such as environmental exposures and healthcare utilization remains possible. Second, despite significant software optimization, analysis of extremely large datasets (e.g., national registries including tens of millions of individuals and extensive numbers of medical conditions) could still be challenging in terms of computational resources, particularly memory capacity. In future developments, we plan to optimize data structures and incorporate data streaming and database-integrated solutions to efficiently manage truly massive EHR data. Third, the analytic performance of ***DiNetxify*** is less optimal when applied to smaller datasets due to limited statistical power. Therefore, we recommend using cohorts with at least 10,000 exposed individuals to reliably perform comprehensive 3D disease network analyses. Finally, it may be important to stress that, in cohorts with exposed and unexposed groups, the workflow characterizes networks and trajectories in aggregate and flags conditions with excess incidence among the exposed. It does not by itself test whether specific networks or trajectories are unique to or amplified in the exposed group, which requires separate analyses of the unexposed group or apply alternative methods tailored to this purpose.

## Conclusion

In this study, we introduced ***DiNetxify***, an innovative, open-source Python package designed to facilitate comprehensive 3D disease network analyses using large-scale EHR data. With its dedicated data class structure, modular design, multiprocessing support, and algorithmic optimizations, ***DiNetxify*** efficiently processes raw EHR records to construct disease networks and visualize the results. Demonstrated through two case studies using UK Biobank data including over half a million individuals, ***DiNetxify*** effectively revealed meaningful disease associations, progression patterns, and clusters aligning with known medical knowledge, while also providing novel insights. By substantially lowering technical barriers with its user-friendly design and flexibility, ***DiNetxify*** has the potential to significantly enhance the adoption of advanced disease network analysis methods among epidemiological and clinical researchers, facilitating deeper exploration and improved understanding of holistic health dynamics from EHR datasets.

## Data Availability

Data from the UK Biobank (http://www.ukbiobank.ac.uk/) are available to all researchers upon making an application.

## Disclosures

The authors report no financial relationships with commercial interests.

## Acknowledgments

The authors thank the team members and colleagues involved in the West China Biomedical Big Data Center-UK Biobank project for their support. Part of this research was conducted using the UK Biobank research resource (application 54803). This work uses data provided by patients and collected by the NHS as part of their care and support. This research used data assets made available by National Safe Haven as part of the Data and Connectivity National Core Study, led by Health Data Research UK in partnership with the Office for National Statistics and funded by UK Research and Innovation.

## Authors’ Contributions

CH and HS were responsible for the study’s concept and design. CH, HL and EUG did the method development. CH and HY did the data cleaning and analysis. CH, HL, VHA, EUG, UAV, YY, FF and HS interpreted the data. CH, HL and HS drafted the manuscript. VHA, EUG, YY, UAV and FF critically revised the manuscript. All the authors approved the final manuscript as submitted and agree to be accountable for all aspects of the work.

## Funding

This work was supported by National Natural Science Foundation of China (grant 82471535 to Dr. Song and 82404391 to Dr. Hou); 1.3.5 Project for Disciplines of Excellence, West China Hospital, Sichuan University (grant ZYYC21005 to Dr. Song); the University Cooperation Grant from NordForsk (PreciMent grant no. 164218 to Drs. Fang and Unnur).

## Ethics approval and consent to participate

The UK Biobank study has received full ethical approval from the NHS National Research Ethics Service (16/NW/0274), and all the participants provided written informed consent before data collection. The current study was approved by the biomedical research ethics committee of West China Hospital (2020.661). All methods were performed in accordance with the relevant guidelines and regulations.

## Code availability

The Python code for the ***DiNetxify*** package and for performing the two case studies is available on GitHub page (https://github.com/HZcohort/DiNetxify).

## Notes

### Competing Interest Statement

The authors have declared no competing interest.

### Author Declarations

The current study was approved by the biomedical research ethics committee of West China Hospital (2020.661).

