## Supplementary Figure for "DiNetxify: a Python package for three-dimensional disease network analysis based on electronic health record data"

### Supplementary Figures

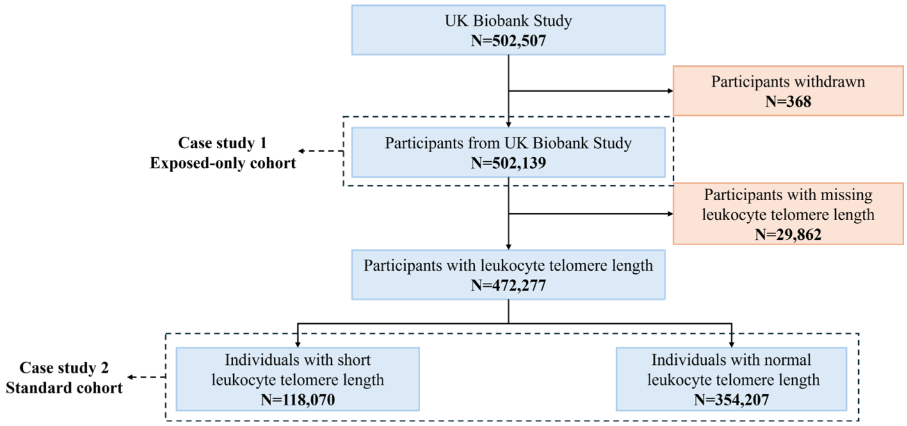


**Supplementary Figure 1** Flowchart for the two case studies using the UK Biobank data

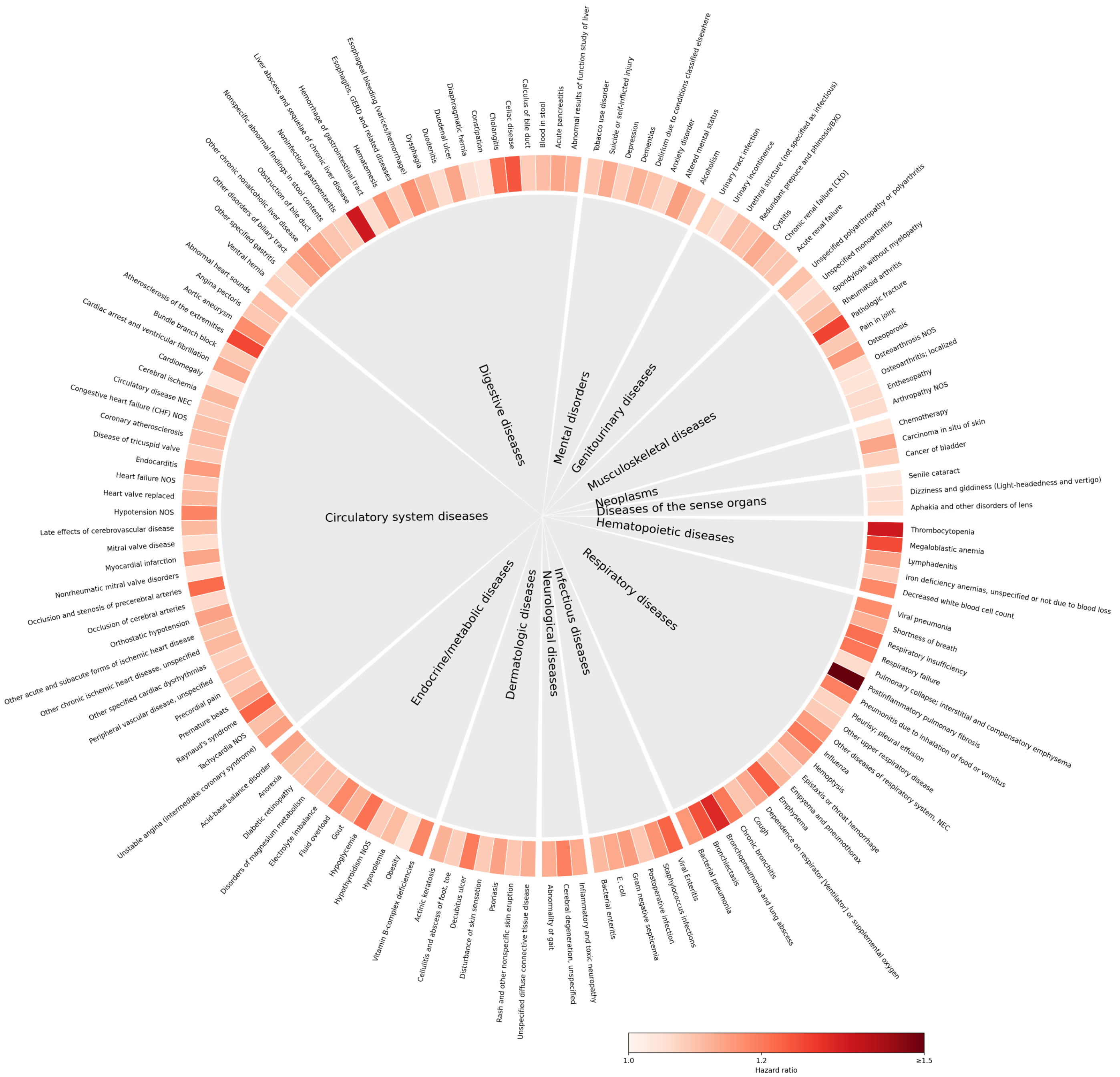


**Supplementary Figure 2** PheWAS analysis results in case study 1.

The ring shows a Hazard Ratio for medical conditions in case study 1 with color intensity reflecting the magnitude of the Hazard Ratio.

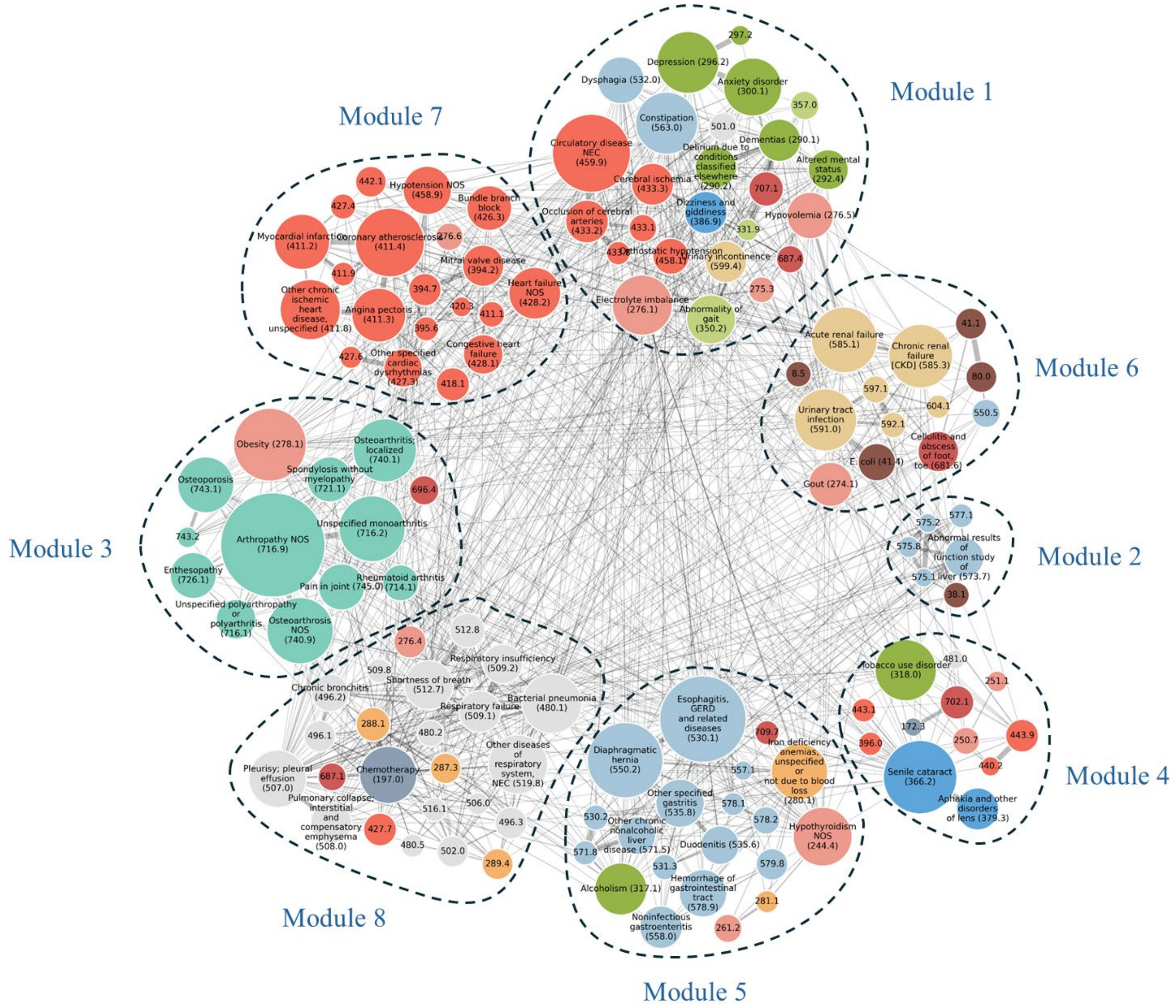


**Supplementary Figure 3** Comorbidity networks following short leukocyte telomere length in case study 1 based on the original regularized partial correlation approach.

Each node represents a disease, where node size indicates prevalence and node color represents the disease category. Both the Louvain and Greedy algorithms partitioned the network into eight modules.

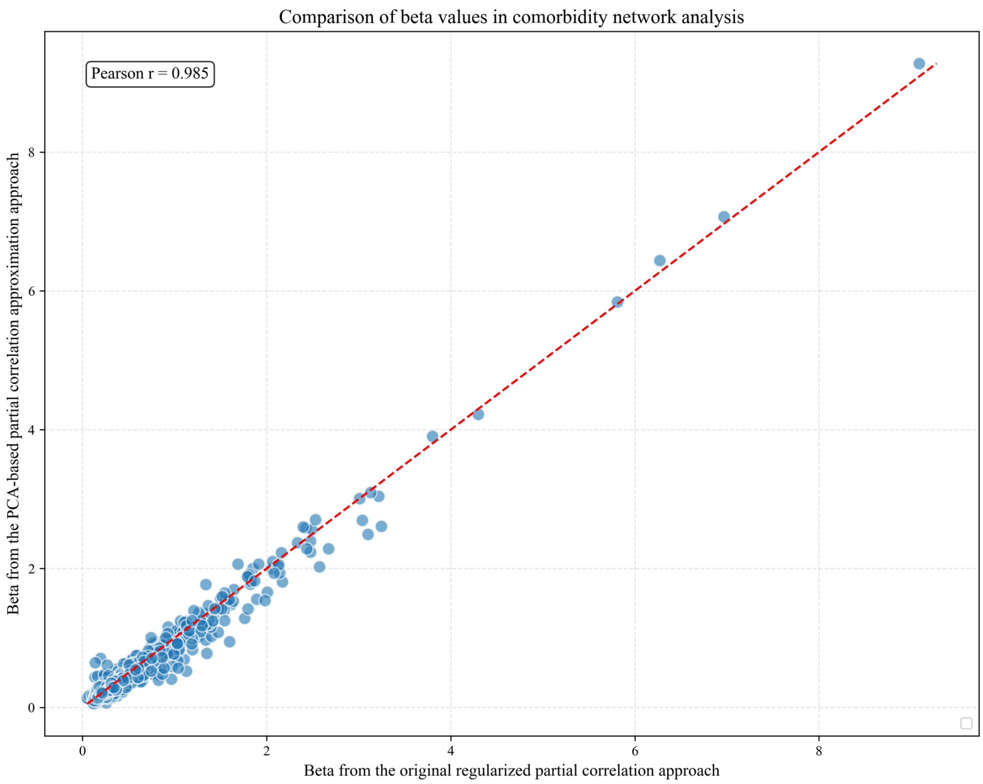


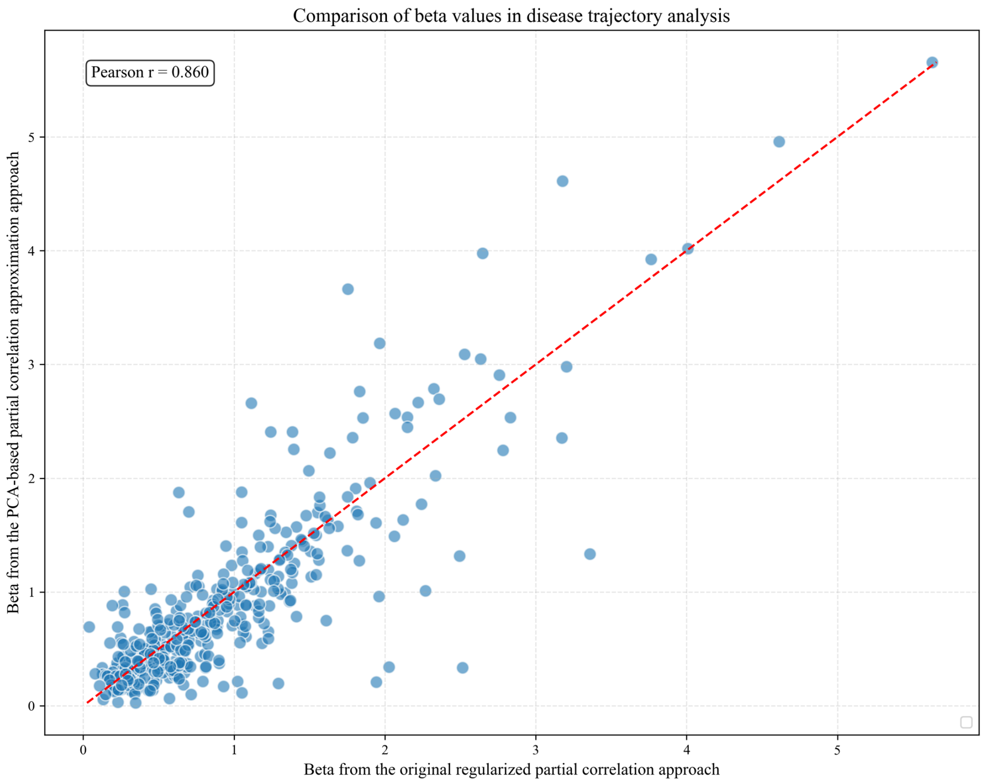


**Supplementary Figure 4** Comparative analysis of two approaches in case study 1.

The top one is the correlation of beta in the comorbidity network analysis from two approaches. The bottom one is correlation of beta in the disease trajectory analysis from two approaches.

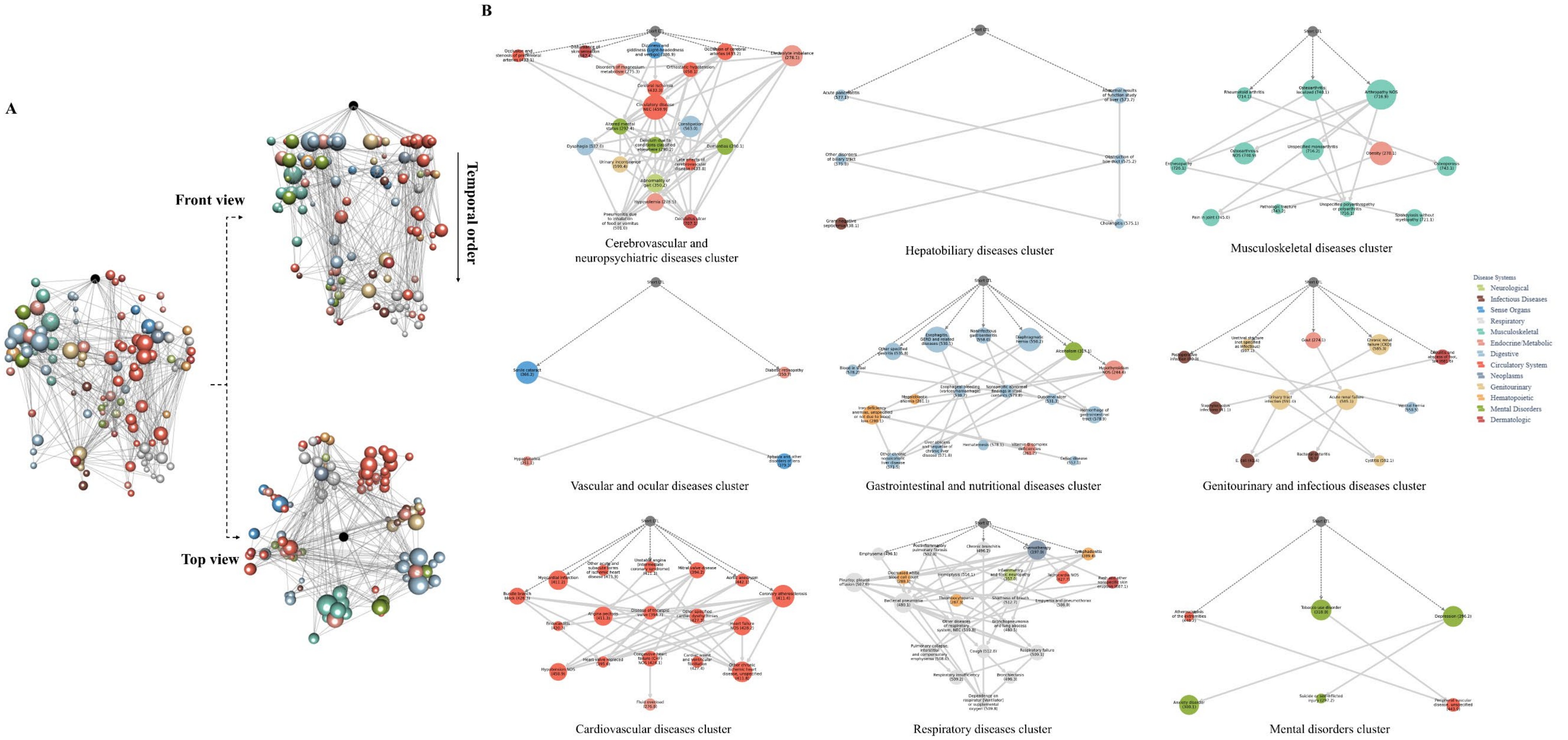


**Supplementary Figure 5** Three-dimensional disease network and the reliable disease clusters associated with short leukocyte telomere length (from PCA-based approximation approach).

Visualization of the disease network associated with short leukocyte telomere length. Nodes represent medical conditions defined by level 2 ‘phecodes’, sized by incidence and colored by disease category. (A) Integrated three-dimensional network plot combining the comorbidity network (viewed from above) and temporal disease trajectory network (viewed from the side), interactive version at https://hzcohort.github.io/DiNetxify/LTL_PCA.html. (B) Nine robust disease clusters revealed through both comorbidity network and disease trajectory analyses.

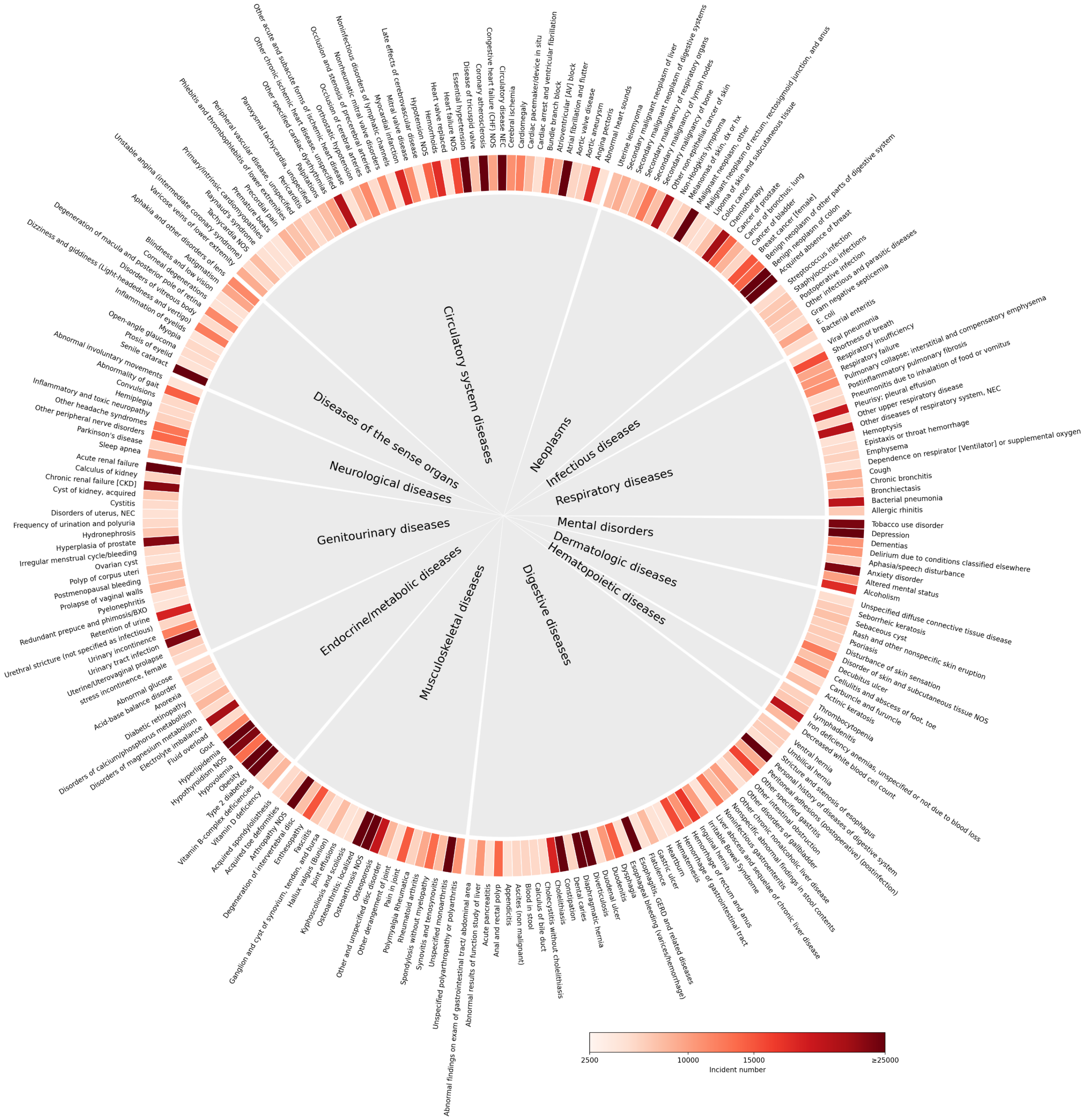


**Supplementary Figure 6** PheWAS analysis results in case study 2.

The ring shows an incident number for medical conditions in case study 2 with color intensity reflecting the magnitude of the incident number.

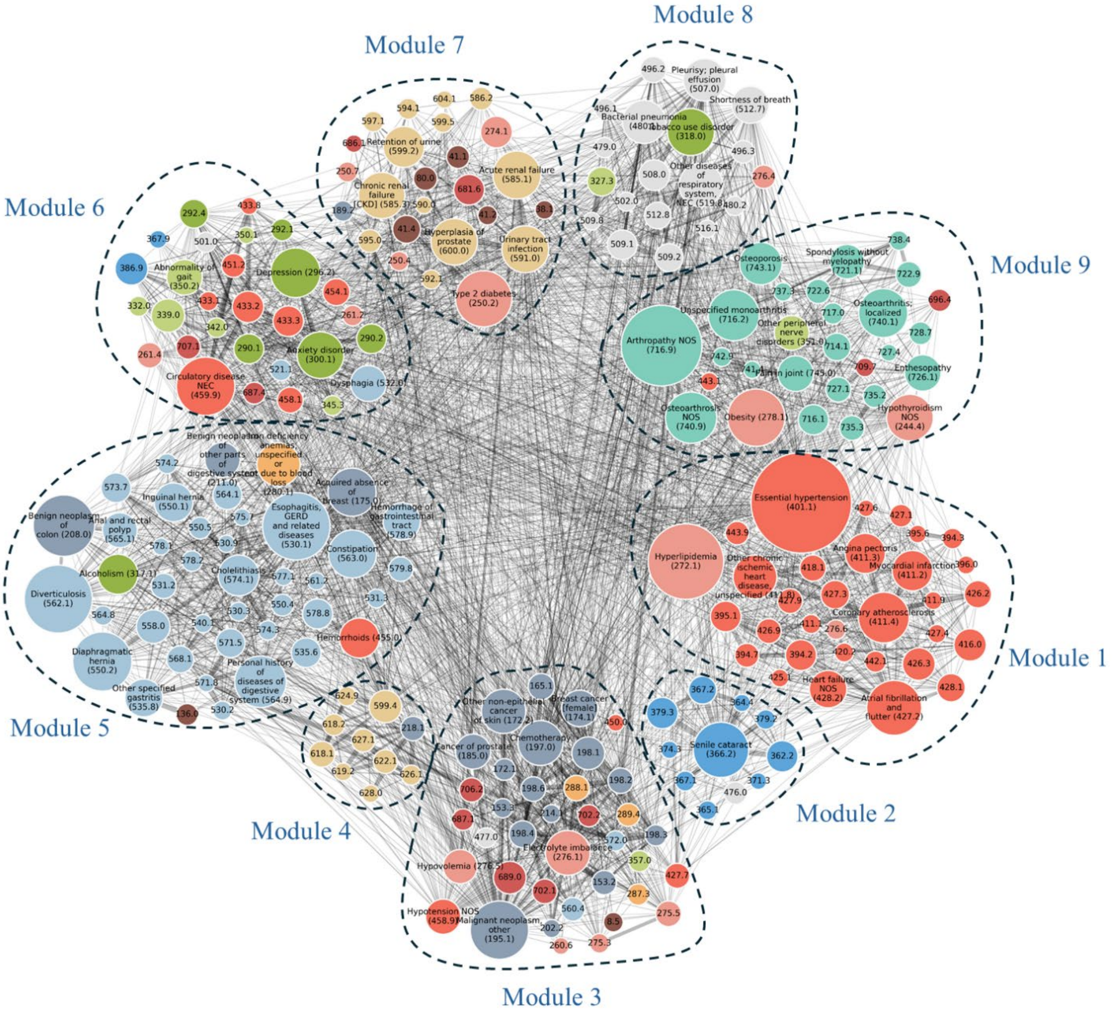


**Supplementary Figure 7** Comorbidity networks in case study 2 based on the PCA-based partial correlation approximation approach.

Each node represents a disease, where node size indicates prevalence and node color represents the disease category. Both the Louvain and Greedy algorithms partitioned the network into nine modules.
